# Prostate cancer screening in hospitalized patients: Results from the Nationwide Inpatient Sample

**DOI:** 10.1101/2021.02.15.21251792

**Authors:** Furqan B. Irfan, Fahad Shabbir Ahmed, Monica V. Masucci, Angelina A. Cerimele, Thu Nguyen, Aryana Sharrak, David M Nguyen, Tooba Tariq, Hussein Baydoun, Ahmad Hasan, Mohamed E Awad, Khaled Saleh, Patrick Karabon

## Abstract

**Background:** Prostate cancer (PCa) is the most common non-skin cancer in American men. The aim of the study was to determine the nationwide prevalence, trends, and predictors of inpatient PCa screening encounters in patients with average risk of PCa using the National Inpatient Sample (NIS) database.

**Methods:** The NIS database from 2006 to 2014 was used to evaluate PCa screening among hospitalized patients in the United States (US). All hospitalized male patients between the ages of 45 and 69 at average risk for PCa were included. The outcome was whether a patient had an encounter for prostate cancer as noted on their discharge record. Variables analyzed included demographic factors, hospital characteristics, and other concomitant diagnoses for prostate or male urinary problems.

**Results:** The prevalence of inpatient PCa screening was 2.57 per 100,000 hospital discharges. In a multivariate setting, the following were significant factors associated with greater odds of inpatient PCa screening: Medicare (AOR: 3.07; P = 0.0016), Self-Pay or Uninsured patients (AOR: 1.74; P = 0.0371), rural (AOR: 11.9; P = < 0.0001) or urban nonteaching hospitals (AOR: 5.26; P = < 0.0001), receiving care in the Midwest (AOR: 4.90; P = < 0.0001), a diagnosis for urinary tract infections (P = 0.0367), genitourinary symptoms (P < 0.0001), hyperplasia of prostate (P = 0.0006), or other male genital disorder (P < 0.0001).

**Conclusion:** According to current cancer screening guidelines, PSA screening should include shared decision making between physicians and patients. In light of unequal access to quality healthcare, there exist disparities in uninsured and rural patients for cancer screening. Screening tools such as prostate specific antigen (PSA) are minimally invasive modalities in the inpatient setting that can help screen individuals at increased risk for the development of prostate cancer, allowing for early detection, prevention, improved rates of cure and ultimately, decreased rates of mortality.

## INTRODUCTION

In the United States, prostate cancer is a leading cause of cancer-related mortality in men. According to the American Cancer Society (ACS), an estimated 248,530 new cases of prostate cancer (PCa) will occur in 2021, with PCa representing one in every five new diagnoses of cancer in men (1). For men, PCa remains the leading cause of new cancer cases and the second leading cause of cancer deaths (2). Since the implementation of prostate specific antigen (PSA) testing in the 1980s, there has been much dispute over the clinical utility of this screening modality due to its potential for overdiagnosis and overtreatment of clinically insignificant disease. Nevertheless, clinicians have observed a notably decreased incidence of distant-stage PCa diagnoses (3). In addition, from 1993 to 2015, the rate of PCa mortality decreased by 52%, which was attributed to early detection via PSA screening and advances in treatment (4). In recent years, however, the death rate is no longer on a downward trajectory.

In 2012, the United States Preventive Services Taskforce (USPSTF) assigned PSA screening a Grade D recommendation, and as a result the overall screening rates declined for all races (3). In particular, the Centers for Disease Control and Prevention (CDC) estimates that AA men have twice as much mortality from PCa when compared with White men. (5) Studies have shown that there are significant racial differences in PCa screening, AA men have decreased rates of PSA screening when compared to Non-Hispanic White men. (6).

In 2018, the USPSTF revised their PSA screening recommendations from a Grade D to a Grade C for men aged 55-69 years. The guidelines suggest that the decision to undergo PSA testing should be a shared one and should be limited to patients who have an interest in undergoing screening (7). However, many patients do not have access to routine health care and preventative screening. In light of these current recommendations, perhaps a more effective screening strategy might meet patients where they are at, wherever they may be interacting with the healthcare system. Whether in an outpatient clinic or admitted to the hospital, physicians can educate and recommend screening in patients they discern are at an increased risk of PCa. Overall, this study aims to determine the nationwide prevalence, trends, and sociodemographic predictors of PCa screening among hospitalized patients with an average-risk of PCa, using data from the National Inpatient Sample (NIS) from 2006-2014.

## METHODS

This population-based, retrospective observational study extracted data from the US National Inpatient Sample (NIS) database. The NIS database was utilized from 2006-2014 to determine the rate of inpatient PCa screening encounters in the USA (8) (9). International Classification of Diseases, 9th revision, Clinical Modification (ICD-9-CM) codes were used to identify and classify hospitalized patients with average-risk of PCa. The ICD-9-CM diagnosis code V76.44 was used to identify inpatients with an encounter for PCa screening. All male inpatients between ages 45 to 69 years in accordance with the most recent USPSTF recommendations (7), that received PCa screening during the study period were included in the study. Inpatients with an increased risk of PCa were then excluded: AA race or the following ICD-9-CM codes: history of PCa (V10.46), benign neoplasm of the prostate (222.2), family history of PCa (V16.42), and genetic susceptibility to malignant neoplasm of prostate (V84.03) (Lynch syndrome, BRCA1&2 mutations, HOXB13 mutation, etc.).

To date, no study has been successful in demonstrating a causal relationship between the diagnosis of an enlarged prostate and the eventual development of PCa. Some epidemiological studies have posited an increased risk of PCa in men with benign prostatic hyperplasia, but these studies lack heterogenous and sizable cohorts of men (10) (11) (12). As a result, we did not exclude patients with benign prostatic hyperplasia from our average-risk group. Patients with malignant neoplasm of prostate or carcinoma in situ of prostate also were included in the study because it is unknown whether the PCa was diagnosed prior to or during the course of hospitalization.

We collected data on patients’ year of hospital admission, age, race, median household income quartile, source of primary payment (patient insurance status), Charlson-Deyo Score (comorbidity index), and hospital characteristics (urban or rural, teaching or nonteaching, bed size). Concomitant diagnoses included: urinary tract infection, genitourinary symptoms, hyperplasia of prostate, and other male genital disorders identified by (ICD-9-CM) codes. The primary outcome was to estimate the prevalence of inpatient PCa screening encounters among hospitalized patients, while secondary outcomes were to determine associated predictors of inpatient PCa screening encounters.

### Statistical analysis

NIS trend weights were utilized to generate nationwide weighted estimates. Variance estimation (strata, clustering, and domain analysis) were carried out according to recommended guidelines (8)(9). Univariate analysis was performed with Complex Samples T-Tests and Chi-Square Tests. Complex samples multivariate logistic regression models were used to determine independent predictors of inpatient PCa screening encounters. Univariate and multivariate analysis results were reported as unadjusted odds ratio and adjusted odds ratio, respectively with confidence intervals and P-Values. Statistical analysis was performed using SAS 9.4 (SAS Institute Inc., Cary, NC, USA) and P-Value < 0.05 indicates a statistically significant association.

## RESULTS

Between 2006 and 2014, the NIS reported a total of 333,933,821 hospitalizations. Of these, 34,379,730 were male patients with an average-risk of PCa that were eligible for inclusion. A total of 884 patients had a documented inpatient PCa screening encounter. Thus, the overall frequency of inpatient PCa screening encounters for average-risk hospitalized patients is rare, approximating 2.57 per 100,000 hospitalizations (Table 1). Fig. 1 illustrates the trend of inpatient PCa screening encounters among hospitalized patients during our study period; however, there was not a significant trend observed during the study period (P = 0.65).

**Table 1:**
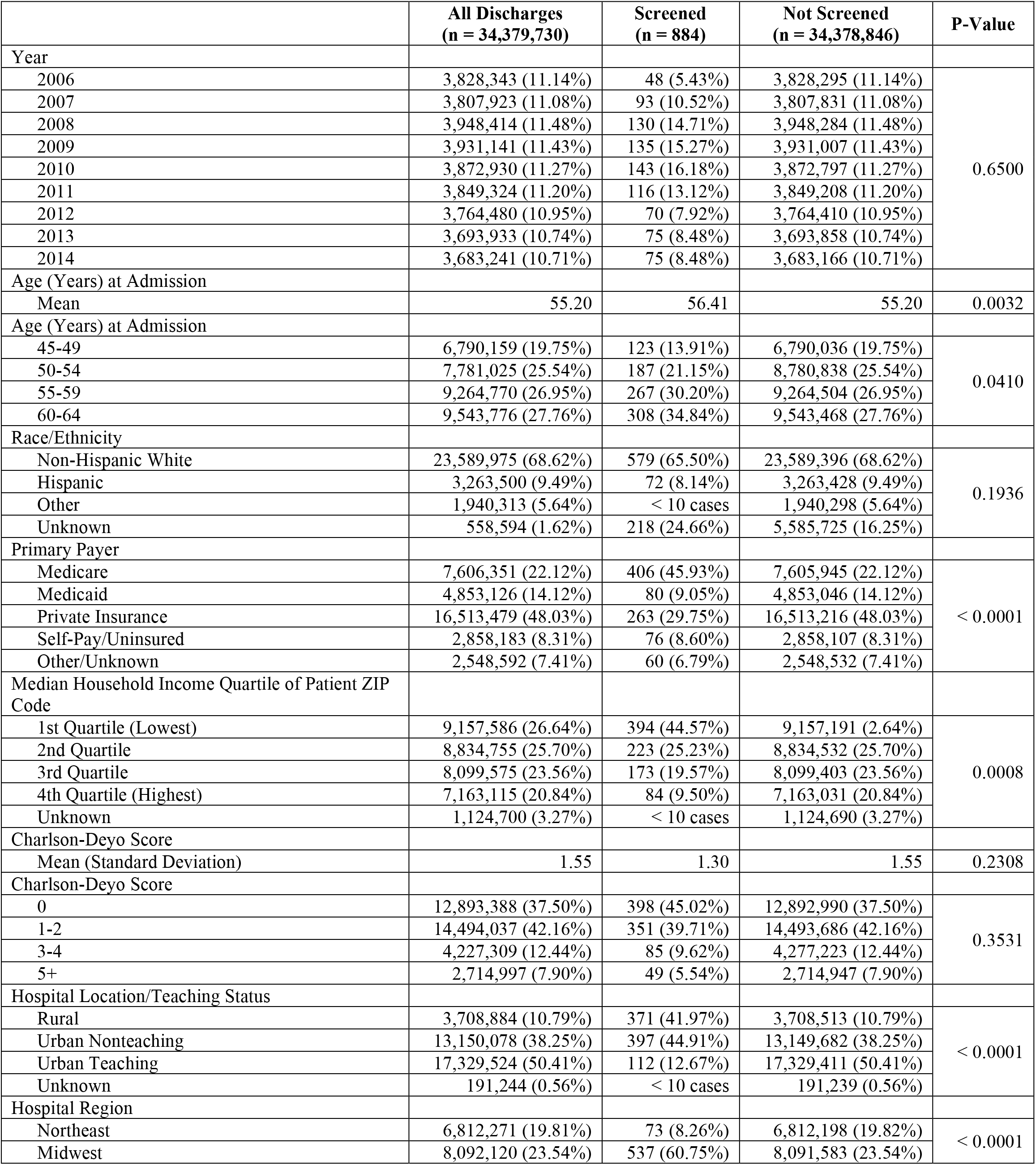
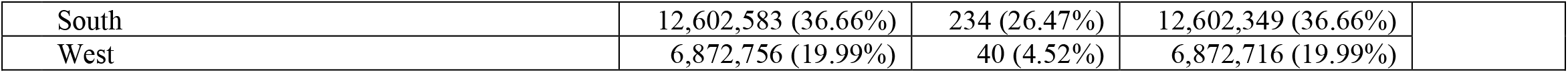
Baseline Demographic and Facility Factors, Stratified by Prostate Cancer Screening Encounter.

|  | **All Discharges**<br>**(n = 34,379,730)** | **Screened**<br>**(n = 884)** | **Not Screened**<br>**(n = 34,378,846)** | **P-Value** |
| --- | --- | --- | --- | --- |
| Year |  |  |  |  |
| 2006 | 3,828,343 (11.14%) | 48 (5.43%) | 3,828,295 (11.14%) | 0.6500 |
| 2007 | 3,807,923 (11.08%) | 93 (10.52%) | 3,807,831 (11.08%) |  |
| 2008 | 3,948,414 (11.48%) | 130 (14.71%) | 3,948,284 (11.48%) |  |
| 2009 | 3,931,141 (11.43%) | 135 (15.27%) | 3,931,007 (11.43%) |  |
| 2010 | 3,872,930 (11.27%) | 143 (16.18%) | 3,872,797 (11.27%) |  |
| 2011 | 3,849,324 (11.20%) | 116 (13.12%) | 3,849,208 (11.20%) |  |
| 2012 | 3,764,480 (10.95%) | 70 (7.92%) | 3,764,410 (10.95%) |  |
| 2013 | 3,693,933 (10.74%) | 75 (8.48%) | 3,693,858 (10.74%) |  |
| 2014 | 3,683,241 (10.71%) | 75 (8.48%) | 3,683,166 (10.71%) |  |
| Age (Years) at Admission |  |  |  |  |
| Mean | 55.20 | 56.41 | 55.20 | 0.0032 |
| Age (Years) at Admission |  |  |  |  |
| 45-49 | 6,790,159 (19.75%) | 123 (13.91%) | 6,790,036 (19.75%) | 0.0410 |
| 50-54 | 7,781,025 (25.54%) | 187 (21.15%) | 8,780,838 (25.54%) |  |
| 55-59 | 9,264,770 (26.95%) | 267 (30.20%) | 9,264,504 (26.95%) |  |
| 60-64 | 9,543,776 (27.76%) | 308 (34.84%) | 9,543,468 (27.76%) |  |
| Race/Ethnicity |  |  |  |  |
| Non-Hispanic White | 23,589,975 (68.62%) | 579 (65.50%) | 23,589,396 (68.62%) | 0.1936 |
| Hispanic | 3,263,500 (9.49%) | 72 (8.14%) | 3,263,428 (9.49%) |  |
| Other | 1,940,313 (5.64%) | < 10 cases | 1,940,298 (5.64%) |  |
| Unknown | 558,594 (1.62%) | 218 (24.66%) | 5,585,725 (16.25%) |  |
| Primary Payer |  |  |  |  |
| Medicare | 7,606,351 (22.12%) | 406 (45.93%) | 7,605,945 (22.12%) | < 0.0001 |
| Medicaid | 4,853,126 (14.12%) | 80 (9.05%) | 4,853,046 (14.12%) |  |
| Private Insurance | 16,513,479 (48.03%) | 263 (29.75%) | 16,513,216 (48.03%) |  |
| Self-Pay/Uninsured | 2,858,183 (8.31%) | 76 (8.60%) | 2,858,107 (8.31%) |  |
| Other/Unknown | 2,548,592 (7.41%) | 60 (6.79%) | 2,548,532 (7.41%) |  |
| Median Household Income Quartile of Patient ZIP Code |  |  |  |  |
| 1st Quartile (Lowest) | 9,157,586 (26.64%) | 394 (44.57%) | 9,157,191 (2.64%) | 0.0008 |
| 2nd Quartile | 8,834,755 (25.70%) | 223 (25.23%) | 8,834,532 (25.70%) |  |
| 3rd Quartile | 8,099,575 (23.56%) | 173 (19.57%) | 8,099,403 (23.56%) |  |
| 4th Quartile (Highest) | 7,163,115 (20.84%) | 84 (9.50%) | 7,163,031 (20.84%) |  |
| Unknown | 1,124,700 (3.27%) | < 10 cases | 1,124,690 (3.27%) |  |
| Charlson-Deyo Score |  |  |  |  |
| Mean (Standard Deviation) | 1.55 | 1.30 | 1.55 | 0.2308 |
| Charlson-Deyo Score |  |  |  |  |
| 0 | 12,893,388 (37.50%) | 398 (45.02%) | 12,892,990 (37.50%) | 0.3531 |
| 1-2 | 14,494,037 (42.16%) | 351 (39.71%) | 14,493,686 (42.16%) |  |
| 3-4 | 4,227,309 (12.44%) | 85 (9.62%) | 4,277,223 (12.44%) |  |
| 5+ | 2,714,997 (7.90%) | 49 (5.54%) | 2,714,947 (7.90%) |  |
| Hospital Location/Teaching Status |  |  |  |  |
| Rural | 3,708,884 (10.79%) | 371 (41.97%) | 3,708,513 (10.79%) | < 0.0001 |
| Urban Nonteaching | 13,150,078 (38.25%) | 397 (44.91%) | 13,149,682 (38.25%) |  |
| Urban Teaching | 17,329,524 (50.41%) | 112 (12.67%) | 17,329,411 (50.41%) |  |
| Unknown | 191,244 (0.56%) | < 10 cases | 191,239 (0.56%) |  |
| Hospital Region |  |  |  |  |
| Northeast | 6,812,271 (19.81%) | 73 (8.26%) | 6,812,198 (19.82%) | < 0.0001 |
| Midwest | 8,092,120 (23.54%) | 537 (60.75%) | 8,091,583 (23.54%) |  |
| South | 12,602,583 (36.66%) | 234 (26.47%) | 12,602,349 (36.66%) |  |
| West | 6,872,756 (19.99%) | 40 (4.52%) | 6,872,716 (19.99%) |  |

**Figure 1:**
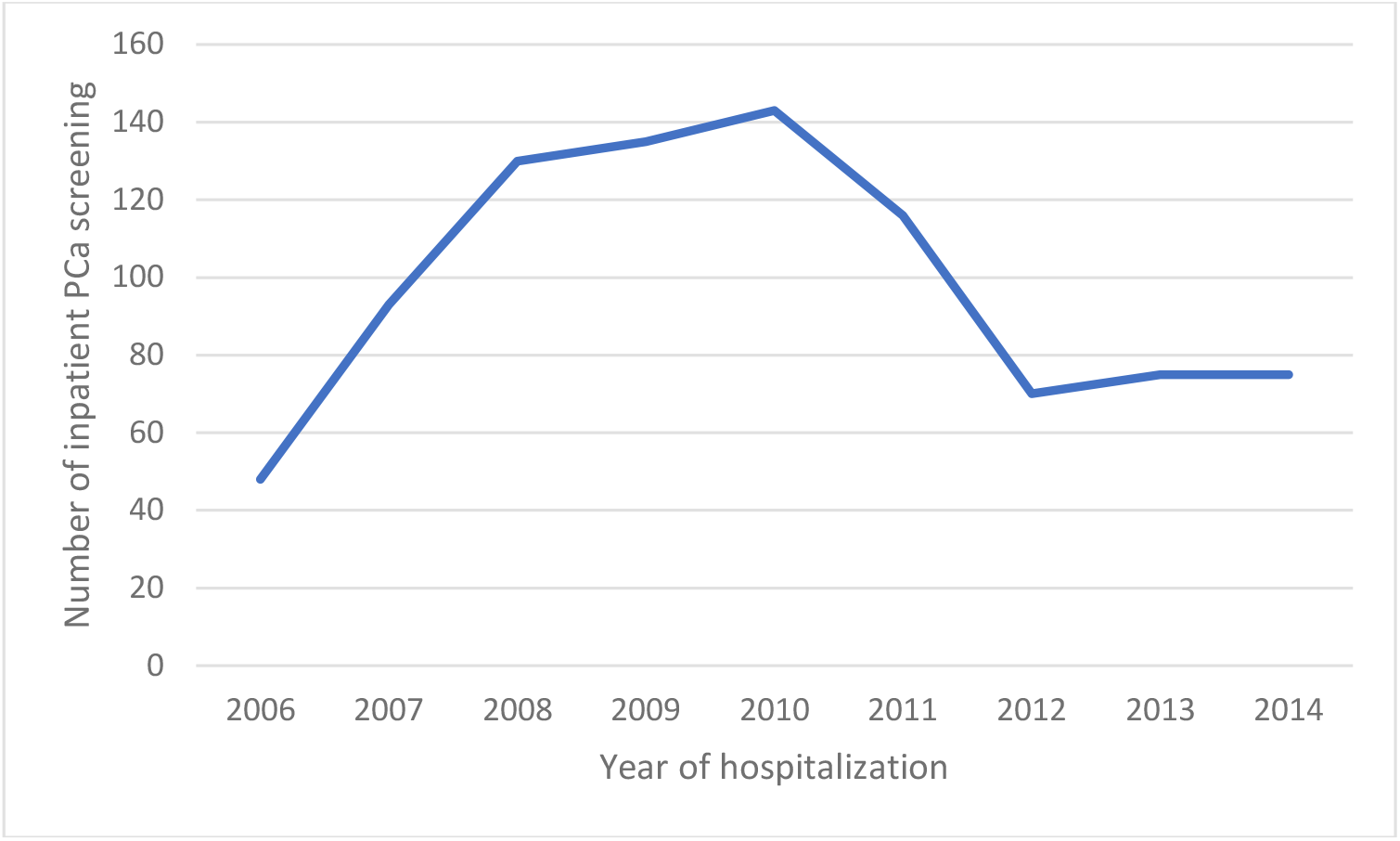
Weighted number of hospitalizations with PCa screening by year of hospital admission.

Factors that could contribute to inpatient PCa screening encounters were evaluated (Table 1). The average age of screened men was 56.41 years. Higher socioeconomic status was associated with lower rates of inpatient PCa screening encounters. The rate went from 4.3 per 100,000 hospitalizations in the lowest quartile of median household income down to 1.17 per 100,000 hospitalizations in the highest quartile of household income (P = 0.0008; Table 1). The rate of PCa screening encounters was much higher in rural hospitals (10/100,000 hospitalizations) than in urban nonteaching (3.02/100,000 hospitalizations), and urban teaching (0.65/100,000 hospitalizations) hospitals (P = < 0.0001). Hospital region also impacted the rate of PCa screening encounters, with the Midwest (60.75%) having a much higher rate of screening than any other region in the country (P = < 0.0001). There was not enough evidence to conclude if there was a significant association between race (P = 0.1936) or Charlson-Deyo Score (P = 0.3531) and PCa screening rates.

Co-occurring diagnoses and procedures associated with PCa screening were isolated (Table 2). Hospitalized men who were screened for PCa were more likely to have a concomitant diagnosis of urinary tract infection, gentourinary symptoms, hyperplasia, and other male genital disorders than those who were not screened. However, it remains unclear whether the diagnosis was made before or after PCa screening was completed.

**Table 2:** Co-Occurring Diagnoses and Procedures, Stratified by Prostate Cancer Screening Encounter.

|  | All Discharges<br>(n = 34,379,730) | Screened<br>(n = 884) | Not Screened<br>(n = 34,378,846) | P-Value |
| --- | --- | --- | --- | --- |
| Urinary Tract Infections<br>Diagnosis |  |  |  |  |
| Yes | 1,503,781 (4.37%) | 69 (7.81%) | 1,503,712 (4.37%) | 0.0367 |
| No | 32,875,949 (95.63%) | 815 (92.19%) | 32,875,134 (95.36%) |  |
| GU Symptoms Diagnosis |  |  |  |  |
| Yes | 1,165,975 (3.39%) | 103 (11.65%) | 1,165,873 (3.39%) | < 0.0001 |
| No | 33,213,755 (96.61%) | 782 (88.46%) | 33,212,973 (96.61%) |  |
| Hyperplasia of Prostate |  |  |  |  |
| Yes | 1,472,653 (4.28%) | 91 (10.29%) | 1,472,562 (4.28%) | 0.0006 |
| No | 32,907,078 (95.72%) | 793 (89.71%) | 32,906,284 (95.72%) |  |
| Other Male Genital Disorders |  |  |  |  |
| Yes | 320,246 (0.93%) | 55 (6.22%) | 320,191 (0.93%) | < 0.0001 |
| No | 34,059,485 (99.07%) | 829 (93.78%) | 34,058,655 (99.07%) |  |

Predictors of PCa screening were weighted in a multivariate logistic regression analysis to determine the factors associated with the observed differences in the rates of screening (Table 3). As compared to men aged 45-69 and adjusted for all other factors in the model, men 55-64 had significantly higher odds of undergoing a PCa screening encounter during hospitalization (all AOR > 1 and P < 0.05). When adjusted for all other factors in the model, Medicare hospitalizations (AOR: 3.07; P = 0.0016) and self-pay/uninsured (AOR: 1.74; P = 0.0371) patients had significantly higher odds of undergoing inpatient PCa screening than private insurance holders. Rural hospitals (AOR: 11.9; P = < 0.0001) and urban Nonteaching hospitals (AOR: 5.26; P = < 0.0001) had significantly higher odds of inpatient PCa screening than urban teaching hospitals when all other factors were adjusted. As compared to South hospitals and adjusted for all other factors in the model, Midwest hospitals had significantly higher odds of inpatient PCa screening encounters (AOR: 4.9; P = < 0.0001).

**Table 3:**
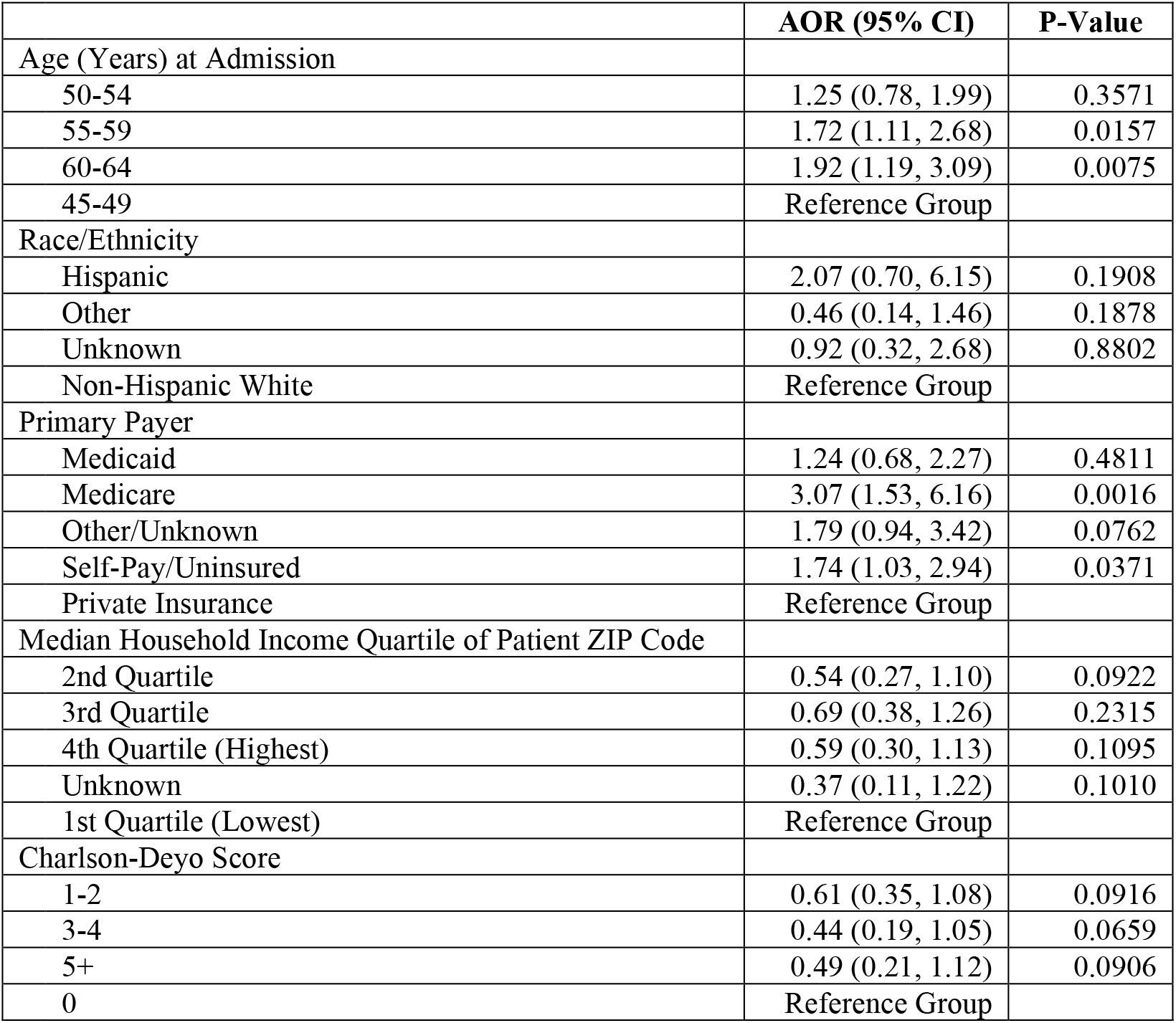
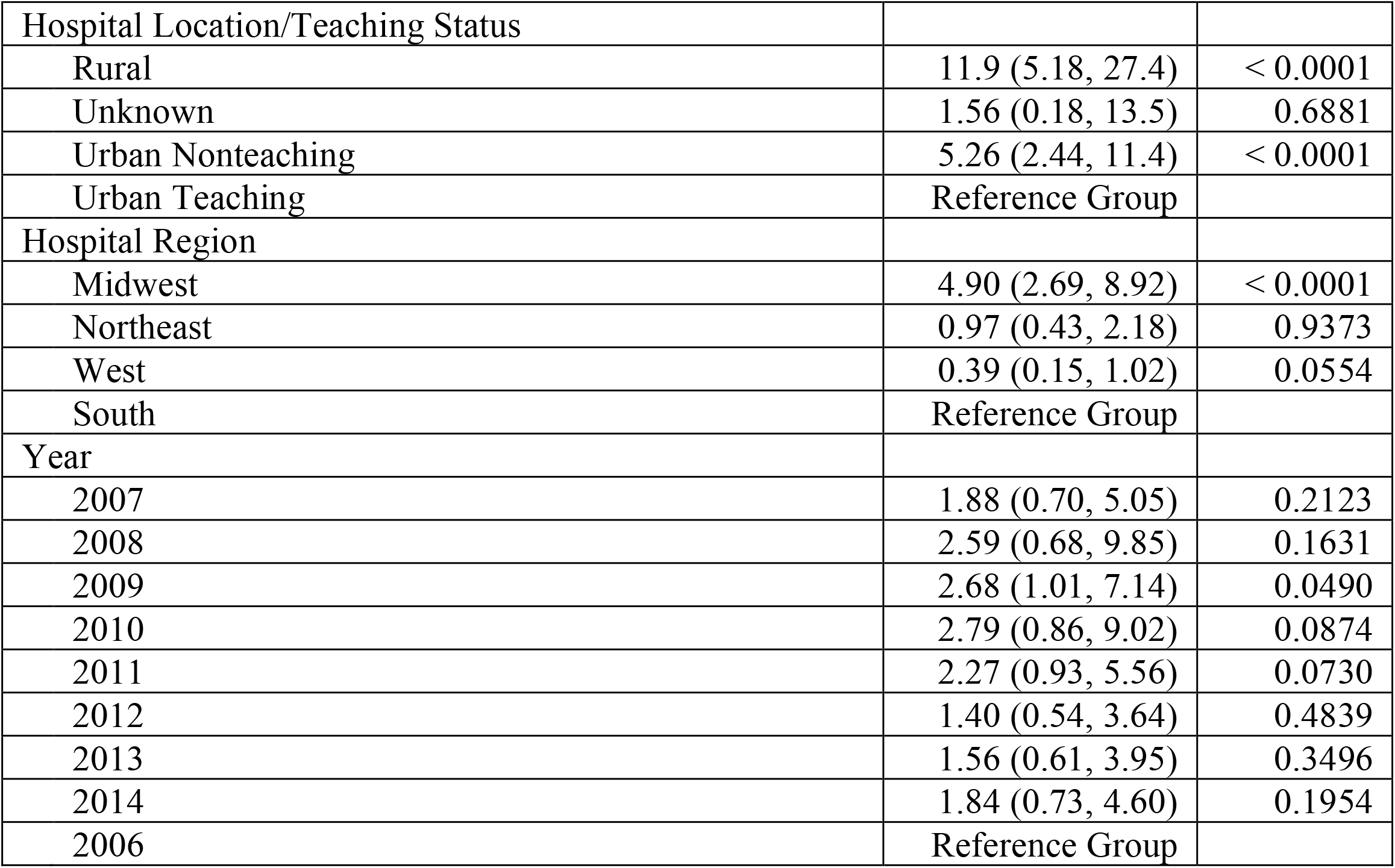
Multivariate Logistic Regression for Prediction of Prostate Cancer Screening Encounter.

## DISCUSSION

This retrospective population-based study’s aim was to determine predictors and trends of inpatient PCa screening. To our knowledge, this is the first population-based study to assess PCa screening in an inpatient population. Our study showed that among 34,379,730 US hospitalizations, only 884 received PCa screening during our eight-year study period. This low rate of inpatient PCa screening is not surprising as patients were likely presenting to the hospital with non-urologic complaints and the primary goals of inpatient care does not necessarily include preventative cancer screenings. In addition, the majority of PCa screening is performed in the outpatient setting, either by a Primary Care Physician (PCP) or Urologist. Discovering an elevated PSA value in the inpatient setting can present its own set of challenges since PSA screening, unlike prostatic biopsy, cannot definitively diagnose PCa and therefore necessitates prompt outpatient follow-up. Aside from potential delays in coordination of care, proper follow-up may be unattainable for many patients as a result of various socioeconomic, geographic, and personal barriers to care. However, if these challenges were successfully mitigated, screening in the inpatient setting could present a promising avenue to provide improved access to cancer screening in populations that have fewer interactions with the healthcare system.

Although our study did not detect significant rates of inpatient PCa screening, outlining the sociodemographic predictors of inpatient PCa screening yielded some noteworthy results. When inpatient PCa screening did occur, it tended to occur in men 55-64 years of age. Hospitalized males with Medicare insurance and those without insurance/self-paid were more likely to have PCa screening. This defines a target population for inpatient cancer screening, represented by those without healthcare insurance who may lack adequate access to healthcare. Patients with private insurance are more likely to have had already been screened or have already had a shared discussion with their PCP or urologist.

Rural hospitalizations were a strong predictor of inpatient cancer screening encounters. Patients admitted in rural health centers may have lower health literacy rates, lack adequate health insurance, have poor healthcare-seeking behavior, and lack adequate access to healthcare (13). For instance, one study found that persons living in rural areas must travel farther distances for preventative breast cancer screenings when compared to their urban area counterparts, and therefore experienced higher rates of late-stage diagnosis of breast cancer (14).

In 2018, the USPSTF changed its 2012 recommendation to a grade “C”. It is thought that this change was likely made in response to the increased rates of and racial disparity in metastatic disease (15). In fact, one study found that the incidence of metastatic PCa at diagnosis increased by about 7% annually from 2007 to 2013 (16). This is particularly concerning since several studies have reported that AA men are at an increased risk for more advanced PCa and higher rates of mortality (17). Other studies project that rates of metastatic PCa will continue to rise considerably in men less than 69 years of age and confirmed that AA over the age of 50 are at two times greater risk of metastatic disease, whereas, AA men younger than 50 are at five times greater risk (18).

As mentioned previously, our study excluded AA men from the dataset as they represent a high-risk profile. Nevertheless, AA men would stand to benefit most from inpatient screening and are at the greatest risk of harm with the newest recommendations to discontinue routine PCa screening. A recent study reiterated the important notion that AA men are more likely to die from PCa and emphasizes the fact that when access to care and treatment is standardized, AA men have comparable mortality outcomes in non-metastatic PCa as their white counterparts (19). With this study’s results in mind, it may be easiest to address the issues of standardized care and access limitations in the hospital setting where resources to do so are more abundant, and thus PCa screening for AA men at risk may be more advantageous. One study reported that “discontinued screening for all men eliminates 100% of overdiagnoses but fails to prevent 100% of avoidable cancer deaths. Continued screening for men aged <70 years eliminates 64% to 66% of overdiagnoses and fails to prevent 36% to 39% of avoidable cancer deaths” (20). It would be beneficial in future research endeavors to delineate whether AA men would benefit more from inpatient screening in comparison to other races, and whether or not they are disproportionately harmed by changing recommendations surrounding PCa screening. This would allow for proper consideration of different patient populations with varying risk profiles and would inform new guidelines that tailor screening protocol according to race and high-risk factors in developing PCa. Given that there was not enough evidence to conclude a significant association between race or Charlson-Deyo Score and PCa screening encounters in our study (P > 0.4905), it could be advantageous to further explore the role of comorbidities among men and if diabetes, hypertension etc., could subsequently increase the need for early PCa screening. Overall, further studies should be conducted that focus on the value of inpatient screening of high-risk males for PCa, in particular, African-American men.

While the NIS database is a uniform, comprehensive, large-scale database that includes all-payers across the United States, the findings of this study are not without limitations, including limitations that are inherent to the database itself. For example, as a result of these limitations, we were unable to distinguish between digital rectal exam (DRE) and PSA testing when the ICD-9-CM code for PCa screening was used. Therefore, we are unable to conclude which type of screening was completed, and thus consistency of screening among different hospitals and patients is unknown. Our study also reports the most common diagnoses in PCa screening encounters included urinary tract infection, genitourinary symptoms, hyperplasia, and other male genital disorders. A third of these diagnoses relate to a urological condition, highlighting potential confounding factors, which may lead to falsely elevated PSA levels. We were unable to determine the timing between concomitant diagnoses during a hospitalization, which created difficulties in deciphering whether the screening encounter occurred before or after a PCa diagnosis when both were listed. The analysis was based on recorded hospitalizations so it is plausible that recorded admissions could have included multiple hospitalizations of a single individual. NIS is publicly available and not updated in real-time with a several year lag in publication of data. Furthermore, it lacks details of outpatient follow-up rates and hospital readmission information since it is restricted to inpatient visits. Lastly, due to its reliance on ICD-9 codes, coding errors remain a possibility along with selection bias (8).

## CONCLUSION

Hospitalized patients represent potential candidates for opportunistic cancer screening of common and preventable malignancies including those of the prostate, colon, breast, lung, and cervix. Our study found that routine inpatient PCa screening encounters in US hospitals was rare. According to current guidelines set by the USPSTF, American Urological Association, American College of Physicians, American Society of Clinical Oncology, American Cancer Society and the National Comprehensive Cancer Network, PSA screening should be a shared decision in patients that indicate a preference (21). In light of the reality that not all Americans have equal access to quality healthcare, this exceptionally low rate of inpatient screening may indicate that there exists a potential opportunity to offer screening to hospitalized patients that may be at increased risk of PCa and who otherwise do not have access to routine cancer screening. Screening tools such as PSA are non-invasive modalities that can help screen individuals at increased risk for the development of prostate cancer, allowing for early detection, prevention, improved rates of cure and ultimately, decreased rates of mortality.

## Data Availability

N/A

